# Epidemiology and genomic analysis of *Klebsiella oxytoca* from a single hospital network in Australia

**DOI:** 10.1101/2022.03.08.22272110

**Authors:** James Stewart, Louise M. Judd, Adam Jenney, Kathryn E. Holt, Kelly L. Wyres, Jane Hawkey

## Abstract

Infections caused by *Klebsiella oxytoca* are the second most common cause of *Klebsiella* infections in humans. Most studies to date have focused on *K. oxytoca* outbreaks and few have examined the broader clinical context of *K. oxytoca*. Here, we collected all clinical isolates identified as *K. oxytoca* in a hospital microbiological diagnostic lab across a 15-month period (n=239). The majority of isolates were sensitive to antimicrobials, however 22 isolates were resistant to third-generation cephalosporins (3GCR), of which five were also carbapenem resistant. Whole genome sequencing of a subset of 92 isolates (all invasive, 3GCR and non-urinary isolates collected >48h after admission) showed those identified as *K. oxytoca* by the clinical laboratory actually encompassed four distinct species (*K. oxytoca, Klebsiella michiganensis, Klebsiella grimontii* and *Klebsiella pasteurii*), referred to as the *K. oxytoca* species complex (KoSC). There was significant diversity within the population, with only 10/67 multi-locus sequence types (STs) represented by more than one isolate. Strain transmission was rare, with only a single likely event identified. Six isolates had either extended spectrum beta-lactamase (*bla*_SHV-12_ and/or *bla*_CTX-M-9_) or carbapenemase (*bla*_IMP-4_) genes. One pair of *K. michiganensis* and *K. pasteurii* genomes carried an identical *bla*_IMP-4_ IncL/M plasmid, indicative of plasmid transmission. Whilst antimicrobial resistance was rare, the resistance plasmids were similar to those found in other *Enterobacterales*, demonstrating that KoSC has access to the same plasmid reservoir and thus there is potential for multi-drug resistance. Further genomic studies are required to improve our understanding of the KoSC population and facilitate investigation into the attributes of successful nosocomial isolates.

## Introduction

The *Klebsiella oxytoca* species complex (KoSC) is a group of Gram-negative bacilli within the order *Enterobacterales* consisting of nine species (1–3) including *Klebsiella michiganensis* (4), *Klebsiella grimontii* (5) and *Klebsiella pasteurii* (6). KoSC are the second most common *Klebsiella* group identified as the cause of clinical infections in humans after the *Klebsiella pneumoniae* species complex (KpSC) (7). Like its sister complex KpSC, KoSC has the ability to persist in a variety of niches, including wet environments (8, 9), hostile environments such as handwashing soaps (10, 11), prosthetic material like central venous catheters (12), and within gastrointestinal flora (13), all of which contribute to its ability to cause opportunistic infections in healthcare settings.

Acquired antimicrobial resistance (AMR) is an emerging concern for KoSC. There are now several reports of multi-drug resistant (MDR) *K. oxytoca* outbreaks carrying either carbapenemases or extended-spectrum beta-lactamases (ESBLs), which confer third-generation cephalosporin resistance (3GCR) (1, 9, 14). In part, this resistance is due to a large shared gene pool with KpSC (1) that can act as a potent source and reservoir of new AMR genes (15). Additionally, 3GCR can emerge through point mutations in the promoter region of the chromosomal *bla*_OXY_ gene (which normally confers only ampicillin resistance), leading to over-production and an ESBL phenotype (16, 17). The most well-characterised virulence factors in KoSC is the kleboxymycin-biosynthetic gene cluster that encodes for the cytotoxin kleboxymcin or tilivallin, both of which are implicated in the pathological changes caused by antibiotic-associated hemorrhagic colitis (3). Most of the remaining virulence information about KoSC has been extrapolated from established *K. pneumoniae* data (1), often without phenotypic evidence.

Despite its clinical relevance and the increasing concerns about AMR, little is known about the epidemiology and broader population structure of KoSC within healthcare institutions. Like KpSC, members of the KoSC remain difficult to distinguish both phenotypically and by matrix-assisted laser desorption/ionization mass spectrometry (MALDI-TOF) (11, 18), and as such are usually reported as *K. oxytoca* by clinical laboratories.

Here, we describe the clinical characteristics of a collection of 239 KoSC infection isolates from hospitalised patients within a single hospital network across a 15-month period. We sequenced a subset of 92 isolates, including all invasive isolates, to characterise their genetic diversity, assess for evidence of nosocomial transmission, and determine the genetic context of acquired AMR determinants.

## Methods

### Ethics

Approval was given by the Alfred Health Hospital Ethics Committee for prospective storage of clinical isolates for future research (HREC 533-16); and later for analysis of the *K. oxytoca* isolates and medical record review for the purpose of this study (HREC 169-19). A consent waiver was granted as the study utilised bacteria that were isolated during routine diagnostic procedures, and hospital records were reviewed only by those who would normally have access to them.

### Specimen collection, identification and antimicrobial susceptibility testing

All KoSC isolated from clinical and screening specimens from 1 January 2018 until 7 April 2019 at the Alfred Hospital Microbiological Diagnostics Laboratory in Melbourne, Australia, were eligible for inclusion (n=239). Bacterial identification was performed using MALDI-TOF (Vitek MS; bioMérieux, Marcy-l’Etoile, France). Antimicrobial susceptibility testing (AST) was performed in the clinical laboratory at the time of isolation using Vitek2 AST-246 (bioMérieux, Marcy-l’Etoile, France), with the exception of urinary isolates where initial disk diffusion susceptibility testing was performed. All MICs and zone of inhibitions were re-interpreted as per EUCAST 2021 guidelines for analysis in this study. Double disk synergy assays were performed later on a subset of isolates to determine which isolates were over-producing their intrinsic *bla*_OXY_ gene.

### Clinical data collection

Clinical information was collected on all 239 isolates from the electronic medical record (EMR) (PowerChart, Cerner, Kansas City, USA). Information regarding microbiological investigations were recovered from the PathNet laboratory information system (LIS) (Cerner, Kansas City, USA). Details on all isolates can be found in **Supplementary Table 1**.

### DNA extraction and sequencing

We selected 92 isolates (38% of the total collection) to sequence using the Illumina platform. This subset included (i) all invasive isolates and all isolates with acquired 3GCR (with the exception of a single isolate that could not be found in storage which was both from a blood culture and had acquired 3GCR, n=39); (ii) all non-urinary isolates collected >48 hours after admission (n=22); (iii) all urinary isolates collected >48 hours after admission with pyuria (WCC>100×10^6^/L, isolated in the absence of co-pathogens or flora, n=6); (iv) all isolates collected during admission to the ICU (n=14); and (v) all non-urinary isolates collected from outpatients or inpatients after <48 hours provided there had been an admission within the prior 12 months (n=11). DNA was extracted from purified cultures using the Agencourt Genfind v2 kit (Beckman Coulter, Beverly, USA) with a paramagnetic based extraction protocol. Library preparation was performed using the Nextera DNA Flex kit (Illumina, San Diego, USA) and libraries were sequenced on the Illumina NextSeq. Six isolates (Kox26, Kox58, Kox71, Kox100, Kox101 and Kox205) were selected for additional long-read sequencing because they carried carbapenemase genes or were involved in suspected transmission events using Nanopore MinION (Oxford Nanopore Technologies, Oxford, UK) with a R9.4.1 flow cell as previously described (19). Whole genome sequencing reads have been deposited under BioProject PRJNA781656 (see **Supplementary Table 1** for individual accessions).

### Genome assembly and pairwise comparisons

To generate draft genome assemblies for genotyping, Illumina reads were assembled using Unicycler v0.4.7 (20) with default parameters. Draft genome assemblies were generated for pairwise single nucleotide variant (SNV) analyses using SKESA v2.2.1 (21) with default parameters, because this comparatively conservative assembly approach removes lower depth sequence regions that are more likely to contain sequence inaccuracies. Catpac (https://github.com/rrwick/Catpac) was used to calculate pairwise SNVs for all pairs of genomes assigned the same ST. SNV counts ≤5 per Mbp were considered indicative of recent transmission, as previously described for the closely related organism *K. pneumoniae* (22).

The six genomes with ONT reads were assembled using Flye v2.8 (23) and polished using Medaka v1.4.3 (https://github.com/nanoporetech/medaka) using the r941_min_hac_g507 model, followed by Illumina polishing using Pilon v1.23 (24). Completed plasmids were compared using BLASTn and comparisons were visualised using genoPlotR v0.8.1 (25). All six completed genome assemblies have been deposited in GenBank, see **Supplementary Table 1** for genome accessions.

### Species confirmation, MLST, AMR, metal resistance and virulence gene screening

Kleborate v2.0.4 (26) was used to identify species, and calculate assembly quality control metrics. Multi-locus sequence types were determined using mlst v2.17 (https://github.com/tseemann/mlst) with the *K. oxytoca* scheme (27).

AMR and metal resistance genes were detected using AMRFinder Plus v3.10.5 (28), specifying species *Klebsiella* and the plus option. Putative pathogenicity and virulence loci were identified through a combination of tBLASTn search for homologues of the key *K. pneumoniae* pathogenicity and virulence determinants using the following loci from the SGH10 reference genome (GenBank accessions CP025080.1 and CP025081.1); SGH10_001765-SGH10_001775 (yersiniabactin, *ybt*, synthesis locus); SGH10_001728-SGH10_001744 (colibactin, *clb*, synthesis locus); SGH10_001661 (*wzi*, capsule synthesis locus marker); SGH10_001680-SGH10_001681 (*wzm* and *wzt* outer lipopolysaccharide synthesis locus markers); SGH10_005357-SGH10_005360 (salmochelin, *iro*, synthesis locus); SGH10_005387-SGH10_005391 (aerobactin, *iuc*, synthesis locus); SGH10_005363 (*rmpA*, regulator of mucoid phenotype gene). tBLASTn hits with ≥70% predicted amino acid identity and ≥95% query coverage were reported (thresholds determined by inspection of the empirical distribution). Additionally, Kaptive v0.7.3 (29) was used to screen for additional evidence of the capsule (K) and outer lipopolysaccharide (O) loci, which were recorded as present if (i) Kaptive identified a match to a locus in the *K. pneumoniae* database with ‘good’ or better confidence; and/or (ii) the *wzi* or *wzm/wzt* marker genes, respectively, were detected by tBLASTn. Putative yersiniabactin locus deletion variants were investigated by manual inspection of the corresponding genome assembly graph in Bandage v0.8.1 (30).

Identification of the kleboxymycin cluster was performed by screening all genome assemblies against the full locus (GenBank accession MF401554) using BLASTn. Genes with >90% coverage and >90% nucleotide identity were considered present. Genomes carrying all 12 genes in the locus were considered to harbour kleboxymycin.

### Core genome phylogeny

All Unicycler assemblies were annotated with Prokka v1.14.6 (31) and a pan-genome was generated using panaroo v1.2.4 (32) with default parameters. A total of 3,502 core genes were detected by panaroo and aligned, the resulting core genome alignment was used to create a core gene phylogeny using IQTree v2 (33) with a GTR+F+I+G4 substitution model. The phylogeny was midpoint rooted and visualised using the R package ggtree (34).

## Results

### KoSC contribute to a diverse range of infections

A total of 239 clinical isolates of KoSC were identified by MALDI-TOF over the 15-month study period, in specimens from 206 individual patients. The patients were 60% male (n=123) with a median age of 67 years (range 20-98) (**Supplementary Table 1**). Specimen types included urine (n=126, 53%), respiratory (n=45, 19%), wound (n=26, 11%), blood (n=19, 8%), tissues and fluids (n=11, 4.6%), and other (n=12, 5%). There were 219 non-sterile specimens with KoSC recovered, of these 51% (n=112) had other microorganisms isolated, 44% of these were urine specimens. Twenty-six patients had multiple isolations of KoSC (median 2, range 2-4). Eighty percent (n=191) of isolates were recovered from inpatients, 38% (n=92) of these were collected >48 hours after admission. Of the 48 isolates collected from outpatients, 58% (n=28) were from individuals with hospital admission or procedures in the prior 12 months. Amongst the 19 isolates recovered from blood, seven were from inpatients >48 hours after admission and nine were from individuals with hospitalisations or procedures in the prior 12 months. The vast majority of isolates were sensitive to antimicrobials except ampicillin, with only 15% (n=22) of isolates displaying a 3GCR phenotype (MIC >2 mg/L to ceftriaxone), of which five were also carbapenem resistant (MIC > 2 mg/L to meropenem).

### Genomic analysis revealed species and substantial sequence type diversity

We sequenced a subset of 92 isolates from the full collection (38%), including all isolates associated with invasive, nosocomial and/or drug-resistant infections (see **Methods**). These isolates represented 31 respiratory specimens (34%), 20 urine (22%), 19 blood (21%), 12 wound/swab (13%), 7 tissue/sterile fluid (7%) and 3 other (3%). We detected four species from the KoSC complex in our collection – 43 *K. oxytoca* (47%), 37 *K. michiganensis* (40%), 8 *K. grimontii* (9%), and 4 *K. pasteurii* (4%) (**Figure 1**). Amongst the 21 invasive isolates (19 from blood, two from tissue/sterile fluid), we found all four species represented – *K. oxytoca* and *K. michiganensis* were the most common (n=10 and n=8 respectively). The remaining three isolates belonged to *K. grimontii* (n=2) and *K. pasteurii* (n=1).

**Figure 1.**
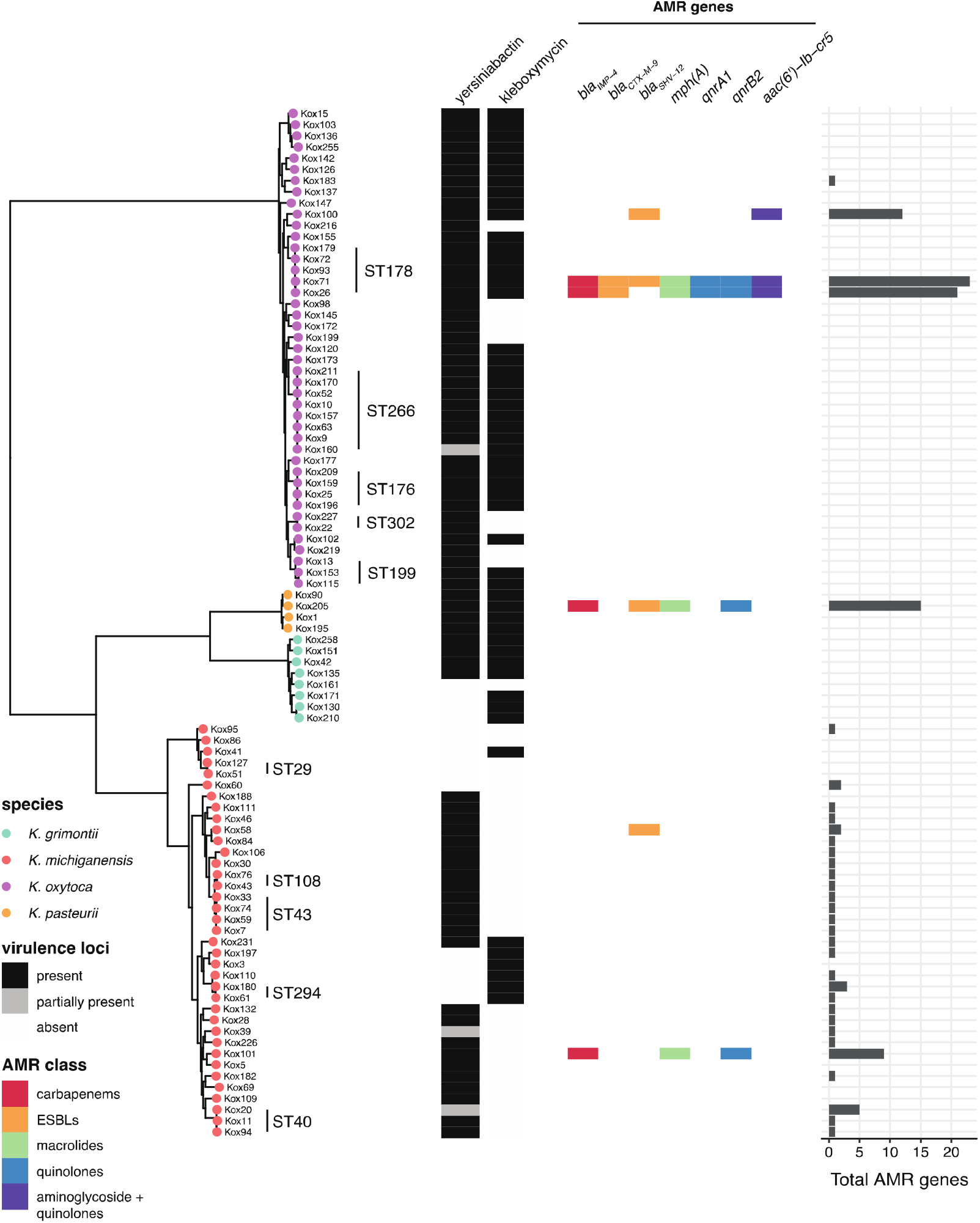
Core genome phylogeny of the KoSC. Left, maximum-likelihood core genome phylogeny (midpoint rooted) of all genomes in this study, with tips coloured by species. STs with ≥2 members are labelled. Heatmap shows presence (coloured) or absence (white) of the virulence loci kleboxymycin and yersiniabactin (as per legend), in addition to key AMR genes (coloured by resistance gene class as per legend). Barchart indicates total number of acquired AMR genes per isolate.

A total of 67 distinct sequence types (STs) were identified, including 43 that were novel to this study. Fifty-seven STs had only a single representative isolate, and a further 10 STs were represented by ≥2 isolates each (**Figure 1**, discussed further below).

### Diversity of resistance, pathogenicity and virulence determinants

The vast majority of genomes (64%, n=59/92) had no acquired AMR genes (**Figure 1**). Of the remaining 33 genomes with at least 1 AMR gene, the majority (n=24, 79%) carried only a chromosomal copy of *aph(3’)-Ia* (which confers aminoglycoside resistance) and no other acquired AMR genes (**Figure 1**). These 24 genomes all belonged to *K. michiganensis*, and overall 70% (n=26) of the *K. michiganensis* population in our study carried a chromosomal *aph(3’)-Ia* gene. Nine genomes carried >1 AMR gene (median 9, range 2-23); six had acquired ESBL and/or carbapenemase genes, accounting for 46% of sequenced 3GCR isolates – one with *bla*_IMP-4_ only (Kox101), two with *bla*_SHV-12_ only (Kox58 and Kox100), one with *bla*_IMP-4_ and *bla*_SHV-12_ (Kox205), one with *bla*_IMP-4_ and *bla*_CTX-M-9_ (Kox26) and one with *bla*_IMP-4_, *bla*_CTX-M-9_ and *bla*_SHV-12_ (Kox71) (**Figure 1**). Of these six isolates, all except Kox58 carried ≥9 AMR genes, conferring resistance to aminoglycosides, macrolides, sulphonamides, tetracyclines, trimethoprim and multiple beta-lactams (**Figure 1, Supplementary Table 1**). Three of the six also carried *mcr9*.*1*, which has been associated with colistin resistance in certain circumstances but was not associated with resistance in these isolates (35). Kox58 was an outlier, carrying only *bla*_SHV-12_ (plasmid-borne) and *aph(3’)-Ia* (chromosomal).

Nine isolates were recognised during phenotypic testing to over-express their chromosomal *bla*_OXY_ gene, eight of these were sequenced (**Supplementary Table 1**). Over-expression leading to 3GCR has been previously associated with point mutations in the *bla*_OXY_ promoter -10 motif at positions 1, 5, 8 and 12, and position 4 in the -35 motif (3, 16, 36). We identified mutations at position 1 (G->T) and position 5 (G->A) of the -10 motif in 12 of our isolates (n=4 G->T and n=8 G->A) (**Supplementary Table 1**); all eight of the isolates identified as over-expressing their *bla*_OXY_ gene during phenotypic testing carried one of these two promotor mutations. However, these mutations were not always associated with 3GCR. Of the eight 3GCR isolates with no acquired ESBL gene detected, six had *bla*_OXY_ promoter mutations but two did not. Additionally, six isolates with *bla*_OXY_ promoter mutations were not 3GCR (MIC range ≤1 - <2 mg/L). Of the five isolates with phenotypic carbapenem resistance, four carried the carbapenemase *bla*_IMP-4_, and one (Kox219) had no known carbapenemase detected. Overall, 86% (n=12/14) of phenotypic 3GCR and 80% of carbapenem resistance in sequenced isolates could be explained by known resistance mechanisms.

The most well-known virulence factor in KoSC is the cytotoxin kleboxymycin. We screened for the full locus (12 genes) required for the production of this toxin (see **Methods**). The locus was present in 56% of genomes (n=52); all *K. pasteurii* genomes (n=4) carried the locus, as well as 87.5% (n=7/8) *K. grimontii* genomes and 79% (n=34/43) of *K. oxytoca* genomes (**Figure 1**). Kleboxymycin was rarer in *K. michiganensis*, present in only 19% (n=7/37) of genomes (**Figure 1**). We also investigated the presence of factors with homology to key virulence determinants recognised for KpSC. We found a K locus (which encodes for the polysaccharide capsule) and an O locus (encoding the outer lipopolysaccharide O antigen) in all isolates, with 35% (n=32/92 of K locus hits and 96% (n=88/92) of O locus hits demonstrating good or very high similarity to previously described KpSC loci (**Supplementary Table 1**). Among the key KpSC acquired virulence loci, only yersiniabactin was detected; the full locus was present in 80% (n=74/92) of genomes, and partially present in 3% (n=3/92) (**Figure 1**). All *K. oxytoca* and *K. pasteurii* genomes carried yersiniabactin, however it was variably present in *K. michiganensis* (70%) and *K. grimontii* (50%) genomes.

### Limited evidence of strain and plasmid transmission

Ten STs (see **Figure 1**) were represented by more than one isolate from either the same or different patients. We conducted a pairwise SNV analysis within each ST to identify putative transmissions between patients (defined as ≤5 SNVs per Mbp, see **Methods**, n=60 pairwise comparisons). The vast majority of pairs (88%, n=53) differed by greater than >100 SNVs (range 5 – 7066). Five pairs differed by 38-104 SNVs (**Table 1**); whilst this does not meet the cut-off for patient-to-patient transmission, it does indicate that these isolates may have been acquired from a common source. Two pairs of isolates differed by ≤30 SNVs. One pair comprised two strains isolated from the same patient 87 days apart which differed by 6 SNVs (Kox170 and Kox211, ST266, **Table 1**). The other pair (Kox26 and Kox71, ST178, **Table 1**) differed by 5 SNVs and were isolated from two different patients 67 days apart, consistent with transmission. These patients were admitted to different wards, and were notable for both carrying a *bla*_IMP-4_ IncHI2A plasmid (see below).

**Table 1:**
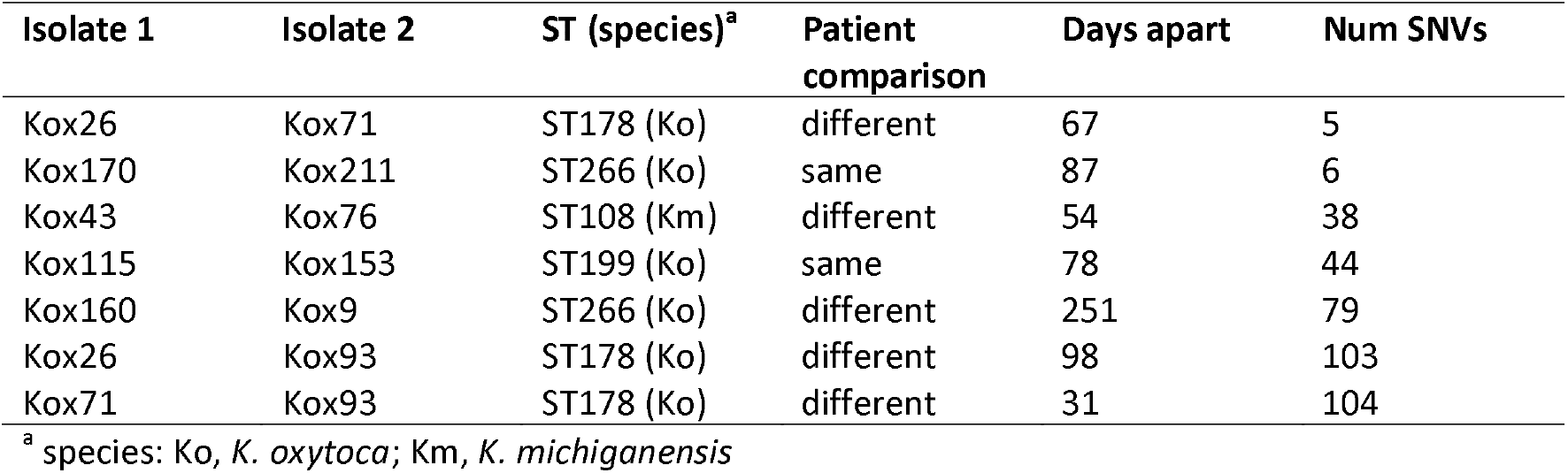
Pairs of isolates differing by <105 SNVs.

In order to determine if there was any evidence of ESBL/carbapenemase plasmid transmission within our hospital we generated completed plasmid sequences for all six isolates that harboured either type of enzyme. We found two distinct plasmids harbouring *bla*_IMP-4_ amongst four genomes (**Figure 2**). Kox101 (*K. michiganensis* ST231) and Kox205 (*K. pasteurii* ST300) carried *bla*_IMP-4_ IncL/M plasmids, which we named pKox101_2 and pKox205_2, respectively (accessions CP089409.1 and CP089405.1). The plasmid sequences were identical in both isolates, suggestive of plasmid transmission between these two species, which were isolated 28 weeks apart from different wards (**Figure 2a**). The *bla*_IMP-4_ IncL/M plasmid carried eight additional AMR genes (*aac(6’)-Ib4, aac(3)-IId, catB3*, two copies of *sul1, qnrB2, mph(A)* and *bla*_TEM-1_), conferring resistance to six different antimicrobial drug classes (aminoglycosides, chloramphenicol, sulphonamides, quinolones, macrolides and penicillins). This *bla*_IMP-4_ IncL/M plasmid is identical to one previously sequenced from an *Enterobacter* genome in Sydney in 2012 (100% nucleotide identity, accession JX101693) (37).

**Figure 2:**
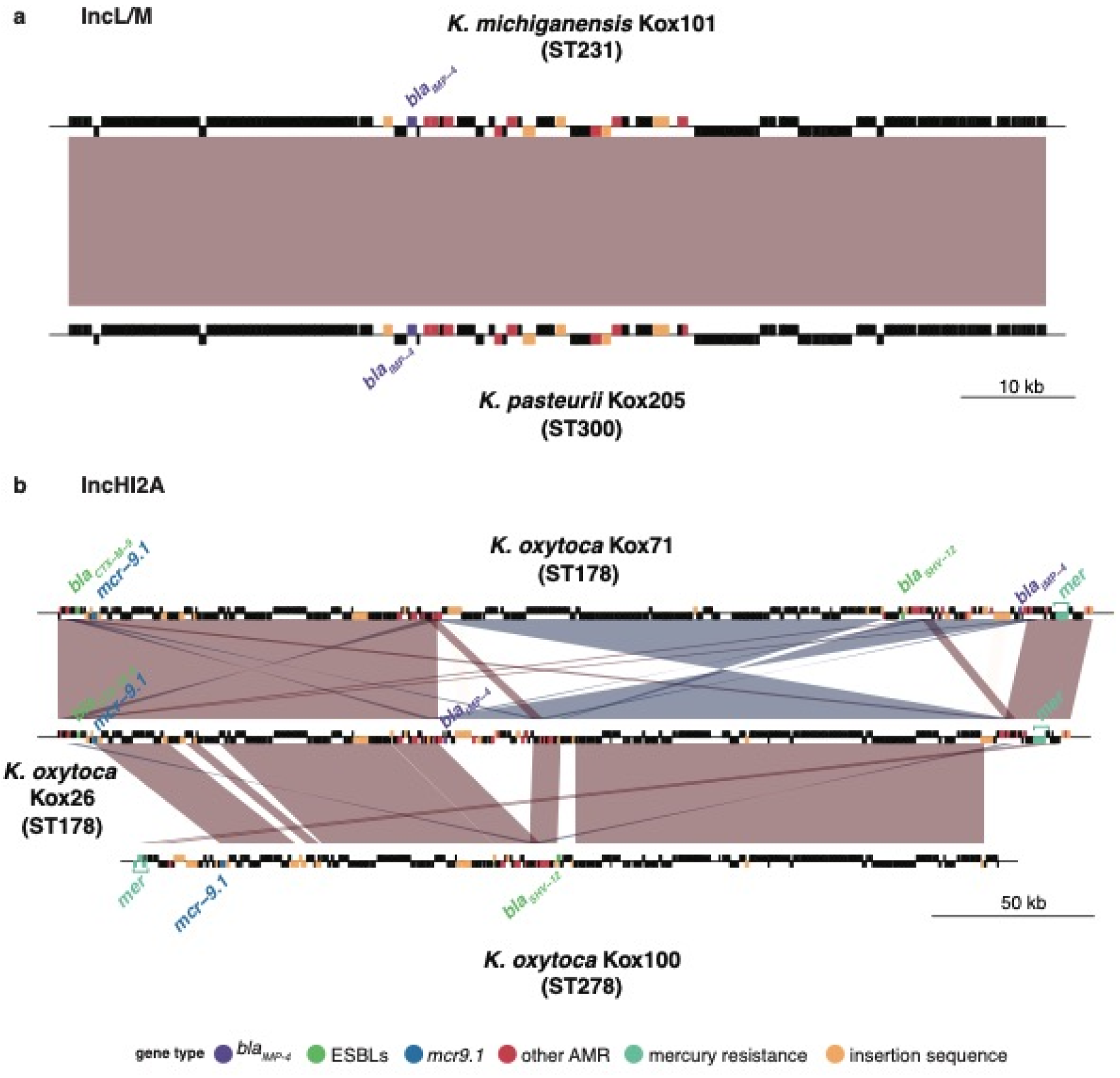
Comparison of carbapenemase and ESBL plasmids. **a)** Comparison of *bla*_IMP-4_ carrying IncL/M plasmids in ST231 and ST300. Blocks indicate genes, top row are genes on forward strand and bottom row are genes on reverse strand. Genes are coloured by type as per legend. Red block indicates >99% nucleotide identity between plasmids. **b)** Comparison of *bla*_IMP-4_ and *bla*_SHV-12_ IncHI2A plasmids in ST178 and ST278. Blocks indicate genes as in a, with genes coloured by type as per legend. Red and blue blocks indicate segments with >99% nucleotide identity between pairs of plasmids, blue indicates the segment is in reverse orientation.

The remaining two *bla*_IMP-4_ positive genomes (both *K. oxytoca*) carried *bla*_IMP-4_ on an IncHI2A plasmid, along with a large number of other AMR genes (n=31 in Kox71, n=28 in Kox26), including the ESBLs *bla*_SHV-12_ and/or *bla*_CTX-M-9_ (**Figure 2b**). Overall, the IncHI2A plasmids conferred resistance to nine antimicrobial drug classes, in addition to heavy metals including mercury. Both plasmids had zero SNVs between them (consistent with strain transmission described above, see **Table 1**), however there has been at least one inversion event and the loss of *bla*_SHV-12_ in Kox26 (**Figure 2b**). A version of this IncHI2A plasmid lacking *bla*_IMP-4_ was detected in Kox100 (ST278), however it was significantly shorter (269 kbp vs ∼300 kbp in ST178), had 212 SNVs in comparison to the ST178 version, and had lost both *bla*_IMP-4_ and *bla*_CTX-M-9_ (though still harboured the ESBL *bla*_SHV-12_, see **Figure 2b**). The Kox100 variant carried 12 additional AMR genes, conferring resistance to eight drug classes. Finally, Kox58 (*K. michiganensis* ST309) carried the ESBL *bla*_SHV-12_ on a 136 kpb IncFII plasmid that harboured silver and copper resistance operons, but zero additional AMR genes.

## Discussion

To our knowledge, this is the first description of the population structure of a non-outbreak collection of isolates identified as *K. oxytoca* from a single hospital network. Isolates from urine were most common, followed by isolates from respiratory and wound specimens, similar to what has been observed for a collection of KpSC infections from the same hospital across a similar time period (38).

We preferentially sequenced hospital onset isolates, invasive isolates and isolates with acquired drug resistance, and found that isolates identified as *K. oxytoca* by the clinical laboratory instead represented four distinct species, with significant diversity within each. Amongst invasive isolates, we found that *K. oxytoca* and *K. michiganensis* were most common, however these species were also the most common across the whole collection. As we did not sequence a representative sample of isolates from all specimen types, we are unable to draw conclusions regarding differences in the rates of urinary tract infections and gastrointestinal carriage for the different species within the KoSC. Improvement in laboratory species identification in the KoSC will be key for bettering our understanding of the clinical syndromes and the likelihood of acquired resistance associated with each species. Models for correctly identifying KoSC species using MALDI-TOF now exist (39, 40), however these models aren’t yet implemented in the MALDI-TOF instrument databases. Whilst spectra can be exported from the instruments and analysed independently, this requires significant additional expertise.

Consistent with earlier studies, we found the virulence factor kleboxymycin gene cluster in all four species in our data, though not conserved in any one species (41). We did not identify any *K. pneumoniae* virulence factors apart from yersiniabactin in our isolate collection, in contrast to some previous studies (1, 42). Given our decision to focus on nosocomial isolates we may have been less likely to identify hypervirulent isolates associated with community-acquired infections, although we did include all invasive isolates with community onset. These findings suggest that these key *K. pneumoniae* virulence factors, apart from yersiniabactin, are uncommon in KoSC. Significantly more work is required to characterise novel virulence factors in KoSC and this will require both phenotypic and genotypic studies.

We observed a marked amount of genomic diversity in our sampled population, collected over a single year from one hospital. The amount of diversity found (where most STs were represented by a single genome) was similar to that found in an earlier study that examined a ten-year collection of *K. oxytoca* genomes from multiple clinical labs in the UK and Ireland (1). Our collection did not include any of the globally distributed STs (ST2 or ST9) (43), or ST315 which was responsible for a *K. michiganensis* outbreak in a neonatal care unit in Australia (11). There was limited evidence of strain transmission in our collection, with only one likely transmission event, of an isolate with an acquired MDR plasmid including *bla*_IMP-4_. However, this event involved patients that were admitted two months apart, and to separate wards, with no overlap in admission times. This may suggest an environmental reservoir or an unrecognised intermediate patient or healthcare worker with carriage or unsampled infection. This low frequency of identified transmission is consistent with previous work in *K. pneumoniae* demonstrating most infections are from a patient’s own flora (22). Previous studies of KoSC have shown close relatedness between environmental and clinical specimens suggesting a possible environmental source for infection (1, 11). We found a number of isolate pairs in our collection with >30 but <105 SNVs, consistent with transmission from an environmental source. Further work sequencing environmental and screening isolates from within the community and hospital will provide context to describe the relative contribution of environmental isolates to carriage and clinical infection and additionally allow for some assessment of candidate virulence factors.

Within our collection AMR was quite rare – only six isolates had acquired multi-drug resistance that included ESBLs. Notably four of these isolates had also acquired the carbapenemase *bla*_IMP-4_. Of these four carbapenemase isolates, two carried an identical IncL/M plasmid that has previously been described in *Enterobacter cloacae* from Sydney (37). The remaining two isolates carried an IncHI2A plasmid that has been found to be circulating in multiple species within our hospital (35), and is highly similar to a plasmid previously described in Brisbane (44), consistent with the previously described widespread dissemination of *bla*_IMP-4_ within the Enterobacteriaceae along Australia’s east coast (45–48). Given that these plasmids are distributed across multiple species in the KoSC and STs, this suggests regular horizontal transfer of plasmids amongst species within this family.

This study provides insight into the clinical epidemiology and population structure of KoSC within a single hospital network, and has identified significant population diversity within what was identified by the clinical laboratory as simply *K. oxytoca. K. oxytoca* was not an uncommon pathogen in our hospital, and whilst AMR was rare, the MDR plasmids identified in our isolates indicate that KoSC has access to the same plasmid reservoir as KpSC, providing the potential for any isolate to become highly drug-resistant with the acquisition of a single plasmid. Future genomic studies will increase our understanding of both community and environmental population structures, which will allow us to further investigate virulence factors and other attributes of successful nosocomial isolates.

## Data Availability

Whole genome sequencing reads have been deposited under BioProject PRJNA781656 (see Supplementary Table 1 for individual accessions).

## Author Contributions

Conceptualization – JS, AJ; Formal analysis – JS, JH, KLW; Funding acquisition – JS, AJ, KEH; Investigation – JS, LMJ, JH, KLW; Resources – JS, AJ, LMJ; Supervision – AJ, JH, KLW; Visualization – JH, KLW; Writing (original draft) – JS; Writing (review and editing) – all authors.

## Supplementary Tables

**Supplementary Table 1: Details of all isolates included in this study**.

